# Self-reported Health Literacy, Digital Literacy, and Barriers to accessing care at a Safety Net Breast Surgical Oncology Clinic

**DOI:** 10.1101/2024.09.20.24314073

**Authors:** Jamie Burke, Madeline G Higgins, Sudheer R Vemuru, Monica Adams, Laura Leonard, Nancy Taft, Gretchen Ahrendt, Kshama Jaiswal, Kristin Rojas, Randy Miles, Sarah E. Tevis

**Affiliations:** University of Colorado School of Medicine, Aurora CO 80304; Department of Surgery, University of Colorado Anschutz Medical Campus, Aurora CO 80304; Adult and Child Center for Health Outcomes Research and Delivery Science (ACCORDS), University of Colorado, Aurora, CO, 80304; Department of Surgery, Denver Health Hospital Authority and Medical Center, Denver CO, 80204; Department of Radiology, Denver Health Hospital Authority and Medical Center, Denver CO, 80204; Department of Surgery, University of Miami Health System, Miami FL 33136

## Abstract

**Background:** Sociodemographic factors, including language, education, race, and insurance status, significantly influence patient outcomes following breast surgery, especially in safety-net hospitals (SNHs) that serve vulnerable populations.

**Objective:** To assess the sociodemographic composition of a breast surgical oncology clinic at an urban SNH and evaluate how these factors impact digital literacy, health literacy, and access to care.

**Methods:** English and Spanish-speaking adult female patients at an urban SNH breast surgical oncology clinic between August and October 2022 completed a survey assessing digital and health literacy, barriers to care, and sociodemographic information. Descriptive statistics and comparative analyses were performed using Chi-squared and Fisher’s exact tests.

**Results:** Of 127 invited patients, 95 (75%) completed the survey. The median age was 50 years. Fifty-six percent identified as Hispanic, 43% preferred Spanish, and 40% had less than a high school education. Health literacy was lower among Spanish-speaking, older, and less-educated patients. Digital literacy was also lower among these groups, with notable disparities in access to computers, the internet, and smartphones.

**Conclusion:** Significant disparities in health and digital literacy exist among vulnerable breast cancer patients at SNHs, particularly among Spanish-speaking, older, and less-educated individuals. Targeted interventions to improve education, access to digital resources, and supportive services are essential to ensure equitable care and improve health outcomes for these populations.

## Introduction

Individual patient characteristics, including demographics^1^, health literacy^2^, and barriers to accessing care^3^, significantly influence patient outcomes following breast surgery. While there has been overall improvement in breast cancer mortality in recent years, this progress is not uniform across different races, ethnicities, socioeconomic groups, and other social demographics^1^. Disparities in outcomes are particularly evident in safety-net hospitals (SNHs) that serve larger numbers of vulnerable populations, such as the uninsured/under-insured, individuals with low socioeconomic status (SES), and communities of color. The individuals of these populations are often affected by risks of poverty, food insecurity, and unstable housing with additional compounded by barriers to accessing medical care such as limited education, language barriers, economic/financial resources, digital literacy, and physical obstacles.^1^. It is crucial to recognize that sociodemographic factors may impact the care received by patients, their disease and medical knowledge, digital literacy, and treatment outcomes^1^. Non-white race and public insurance status (Medicaid or Medicare) are correlated with higher mortality rates, later stage cancer diagnoses, and delays in treatment^1^.

### Health Literacy

Health literacy may also have a profound effect on patient outcomes. According to Cordasco et al., higher education levels correlate with increased rates of survival^4^ whereas low levels of health literacy are associated with increased emergency department revisits^5^, decreased medication and treatment adherence^2^, fewer numbers of mammography screening and vaccinations^2^, and among elderly patients, higher rates of mortality and poorer health status ^2^. Providers often overestimate patient literacy, and due to shame or reluctance, patients may not disclose literacy challenges^6^. Social factors such as education level, income, and ethnicity have been associated with statistically significant lower health literacy ^7^.

### Non-English-Speaking

Similar deficits in treatment adherence, disease knowledge, and patient outcomes are seen among non-English speaking patients, who also incur greater healthcare costs as a result^4^. These greater costs are driven by more frequent visits to physicians, nonphysicians, and emergency departments as well as increased prescription costs related to increased disease severity and inadequate management of prescriptions^8^. Non-English-speaking patients often report lower health literacy and digital literacy, which significantly impacts their disease knowledge and adherence to medical treatments^3^. These patients frequently encounter challenges in understanding medical instructions, accessing health information, and navigating healthcare systems. The language barrier exacerbates these issues, as many health resources are predominantly available in English. Consequently, non-English-speaking patients may have limited comprehension of their diagnosis, treatment options, and medication regimens. This lack of understanding can lead to decreased adherence to prescribed treatments, resulting in poorer health outcomes and increased healthcare utilization^6^.

### Digital Access

Within the SNH population, additional barriers to digital literacy and online portal usage include limited computer/internet access, negative previous experiences, and lack of computer skills^3^. Research by Choi & DiNitto demonstrated that non-White individuals were more likely to have never used the internet with Black and Hispanic individuals nearly 4.5 times more likely to have never used the internet^9^.

Furthermore, for every unit increase in the income-to-needs ratio (the ratio between income to the official poverty line adjusted for the family size), the likelihood of never using the internet decreased by 36%^9^.

### Breast Cancer Population

Over the past several years, breast cancer survival has improved significantly with the relative survival rate approaching 91%^1^; however, this improvement is not shared equally among various races, ethnicities, socioeconomic groups, and other social demographics. Spanish-language-proficient (SLP) experience higher rates of mastectomies, but lower rates of breast reconstruction surgeries^10^.

Additionally Spanish-language-proficient patients report less satisfaction, functional well-being, and emotional well-being with care than English-language-proficient (ELP) patients ^10^. Patient demographics impact the risks that individuals face. Young women of color are generally diagnosed with breast cancers of higher biological risk as compared to European American women^11^. These biological risks include premenopausal, later stage, and more aggressive tumors that are more difficult to treat. In general, racial/ethnic minorities and patients of lower socioeconomic status face greater health care system challenges including delays in diagnostic and therapeutic care, uncoordinated care, poor provider communication, and poor treatment satisfaction ^10^. Factors such as Ethnicity, native language, and insurance status correlate with varying degrees of patient reported outcomes. Non-white, Spanish-speaking, or uninsured patients typically experience higher biological risk cancers, worse patient-reported outcomes, and poor patient-provider communication.^10^.

Breast cancer patients receiving care at SNHs may be adversely affected by the myriad of barriers they encounter while seeking medical treatment. This study aims to assess the sociodemographic composition of a busy breast surgical oncology clinic at an urban SNH and evaluate how language, education, race, and insurance status impact digital literacy, health literacy, and access to care. The goal is to identify areas of improvement to ensure equitable care for SNH patients, enabling them to better understand their disease process and medical care.

## Methods

### Study population

English and Spanish-speaking adult female patients who were seen at a breast surgical oncology clinic at an urban SNH between August and October 2022 were eligible to participate in this study. This study was approved as a quality improvement project by the Quality Improvement Review Committee (QuIRC #382) of Denver Health and Hospital Authority and thus was exempt from institutional review board review.

### Study Design

Eligible patients were approached to complete a paper survey available in both English and Spanish upon checking into their clinic appointment. The survey (Appendix A) included items evaluating digital and health literacy as well as items querying barriers to accessing breast cancer care and sociodemographic information such as: age, education level, preferred language, race, ethnicity, health insurance type, and prior engagement with primary care and specialty care clinicians. Digital and health literacy items were adapted from validated digital and health literacy questionnaires^12,13^. All items in the survey were reviewed and approved by a group of experts in the field of breast surgical oncology as well as staff who were well acquainted with the patient experience and workflow of the clinic. The English language version of the survey was then translated using a professional medical research translation service (MacMillan Language Services Inc; Erie, CO, USA). Completed paper surveys were subsequently uploaded electronically for secure storage and tabulation using the Research Electronic Data Capture (REDCap) electronic data capture tools hosted at the University of Colorado Anschutz^14,15^.

### Statistical analysis

Descriptive statistics for all sociodemographic items were reported. Responses to the digital and health literacy items were compared using Chi-squared and Fisher’s exact tests. All statistical analyses were performed using IBM SPSS Statistics for Macintosh, Version 28.0 (IBM Corp, Armonk, NY, USA). All tests were two-tailed; the threshold for statistical significance was p<0.05.

## Results

### Participant Demographics

Of the 127 patients invited to complete the questionnaire, 95 completed it (75% completion). Demographics reported included age, race, ethnicity, preferred language and interpreter use, as well as insurance status (Table 1). The median age of respondents was 50 years (N=88) with interquartile range of 43-59. Women aged 40-74 consisted of 71.6% of respondents, while women aged <40 years of age 15.8%, and >75 years 5.3%. Fifty-six percent of respondents identified as Hispanic while 24.2% reported their race as white, 16.8% as black, 2.1% as American Indian, 4.2% as Asian or Pacific Islander, 10.5% as other, and 22.1% as no race. Forty-three percent preferred Spanish, while 52.6% preferred English, with a substantial portion (38.9%) requiring a medical interpreter. Forty percent of respondents had less than a high school education. Fifty percent disclosed using public health insurance while 28.4% reported being uninsured. Most patients had an active primary care physician (77.9%) and had seen their primary care physician within the past 18 months (78.9%).

**Table 1.** Patient Demographics.

|  | N (95) | Frequency (%) | Mean | SD |
| --- | --- | --- | --- | --- |
| **Median Age, years (SD)** | 88 |  | 50.0 | 13.94 |
| < 40 years | 15 | 15.8 |  |  |
| 40-74 years | 68 | 71.6 |  |  |
| ≥75 years | 5 | 5.3 |  |  |
| Unknown | 7 | 7.4 |  |  |
| **Race** |  |  |  |  |
| White | 23 | 24.2 |  |  |
| Black or African American | 16 | 16.8 |  |  |
| Asian / Pacific Islander | 4 | 4.2 |  |  |
| Native American or American Indian | 2 | 2.1 |  |  |
| Other | 10 | 10.5 |  |  |
| I do not identify with a racial group | 19 | 20.0 |  |  |
| Unknown | 21 | 22.1 |  |  |
| **Ethnicity** |  |  |  |  |
| Hispanic, Chicano, or Latinx | 54 | 56.8 |  |  |
| Non-Hispanic, Chicano, or Latinx | 34 | 35.8 |  |  |
| **Preferred Language** |  |  |  |  |
| English | 50 | 52.6 |  |  |
| Spanish | 41 | 43.2 |  |  |
| **Need Interpreter** |  |  |  |  |
| Yes | 37 | 38.9 |  |  |
| No | 53 | 55.8 |  |  |
| **Education Level** |  |  |  |  |
| Less than High School | 37 | 38.9 |  |  |
| High School, Some College, Trade School | 37 | 38.9 |  |  |
| College | 16 | 16.8 |  |  |
| Unknown | 5 | 5.3 |  |  |
| **Healthcare worker** |  |  |  |  |
| Yes | 9 | 9.5 |  |  |
| No | 81 | 85.3 |  |  |
| **Health Insurance Status** |  |  |  |  |
| I do not have health insurance | 27 | 28.4 |  |  |
| I have Medicaid | 39 | 41.1 |  |  |
| I have Medicare | 9 | 9.5 |  |  |
| I have private health insurance | 10 | 10.5 |  |  |
| Unknown | 10 | 10.5 |  |  |
| **PCP** |  |  |  |  |
| Yes | 74 | 77.9 |  |  |
| No | 16 | 16.8 |  |  |
| **Seen PCP in the last 18 months** |  |  |  |  |
| Yes | 75 | 78.9 |  |  |
| No | 15 | 15.8 |  |  |
| **Being followed or treated for breast cancer** |  |  |  |  |
| Yes | 56 | 58.9 |  |  |
| No | 32 | 33.7 |  |  |

### Health Literacy

Overall, 75% of patients reported ‘always’, ‘often’ or ‘sometimes’ having someone help them read hospital materials (Figure 1). In the BRIEF health literacy questionnaire (Figure 1), there was lower health literacy among Spanish speaking individuals, older patients, as well as those with lower levels of education. Spanish speaking patients report requiring more help reading hospital material, discomfort filling out medical forms by themselves, and difficulty understanding written information about their medical condition. Nearly half of patients with less than high school education frequently require help with reading hospital material compared to about 10% in those with high school education or more.

**Figure 1.**
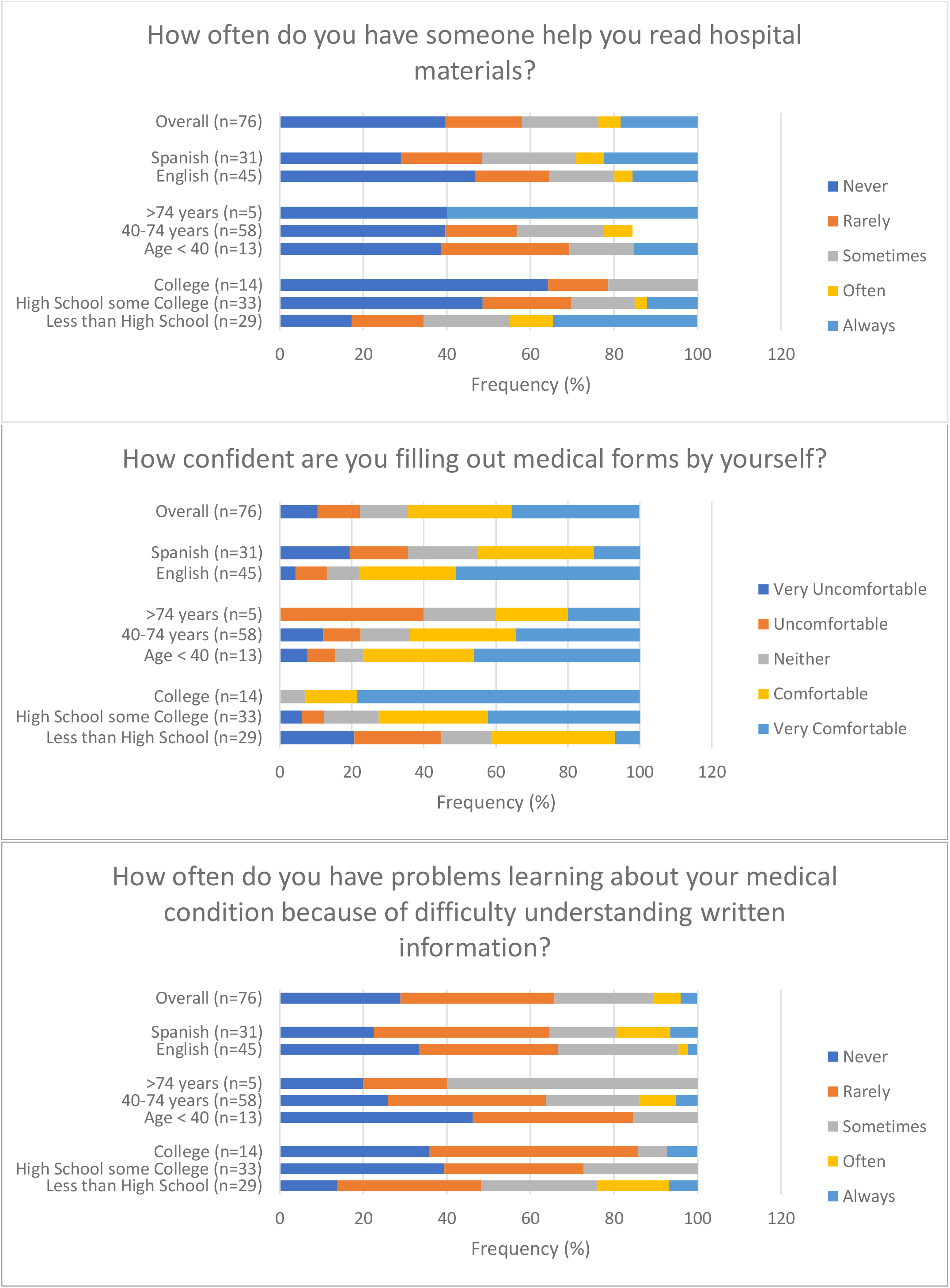
BRIEF Health Literacy Overall.

### Digital Literacy

Computer, internet, and smartphone access was analyzed across age ranges, native language, and education status (Table 2, Figure 2). Increasing in age showed a decrease in computer access from 73% in those aged <40, to 44% in those aged 40-74, and down to 40% for those >75 years old. Internet access showed a similar trend with 93.3% aged <40 years old with access, 77.9% among those aged 40-74, and 60% aged 75 years or older with access to internet. The most drastic change was among smartphone access with 93.3% of those aged <40 years with access to a smartphone, 73.5% among 40–74 years old, and 40% of those aged 75 years or older. Patient’s preferred language compared across digital platforms was also compared. Access to a computer was 64% among English speaking patients and 29.3% among Spanish speaking patients. Eighty-two percent of English-speaking patients reported access to the internet in comparison to 73.2% of Spanish speaking patients. Smartphone access was reported at 78% among English speakers compared to 68.4% among Spanish speakers. Additionally, education status was applied to digital resource access. Among those with less than high school level education, 27% reported access to computers, 70.3% reported access to internet, and 67.6% reported access to a smartphone.

**Table 2.** Frequency of poor digital literacy stratified by education level, age, and preferred language. Poor digital literacy was defined as answer ‘very poor’ or ‘poor’. Missing values were removed for an overall N=83.

| Poor Digital Literacy by Tool | Less than High School<br>N=33 (%) | High School, Some College<br>N=35 (%) | College<br>N=15 (%) | $\chi^2$ (p-value) |
| --- | --- | --- | --- | --- |
| Computer | 28 (84.8) | 6 (17.1) | 1 (6.7) | 42.495 (<0.001) |
| Smartphone | 23 (69.7) | 6 (17.1) | 1 (6.7) | 32.838 (<0.001) |
| Internet | 25 (75.8) | 5 (14.3) | 2 (13.3) | 28.203 (<0.001) |

| Poor Digital Literacy by Tool | Age <40 years<br>N=14 (%) | Age 40-74 years<br>N=64 (%) | Age >74 years<br>N=5 (%) | $\chi^2$ (p-value) |
| --- | --- | --- | --- | --- |
| Computer | 0 (0.0) | 32 (50.0) | 3 (60.0) | 12.470 (0.002)* |
| Smartphone | 1 (7.1) | 26 (40.6) | 3 (60.0) | 6.891 (0.032)* |
| Internet | 1 (7.1) | 28 (43.8) | 3 (60.0) | 7.531 (0.023)* |

\*2 cells have expected count less than 5 due to few patients in the <40 and >74 years old groups.

| Poor Digital Literacy by Tool | English<br>N=47 (%) | Spanish<br>N=36 (%) | $\chi^2$ (p-value) |
| --- | --- | --- | --- |
| Computer | 10 (21.3) | 25 (69.4) | 19.395 (<0.001) |
| Smartphone | 9 (19.1) | 21 (58.3) | 13.561 (<0.001) |
| Internet | 11 (23.4) | 21 (58.3) | 10.499 (<0.001) |

**Figure 2.**
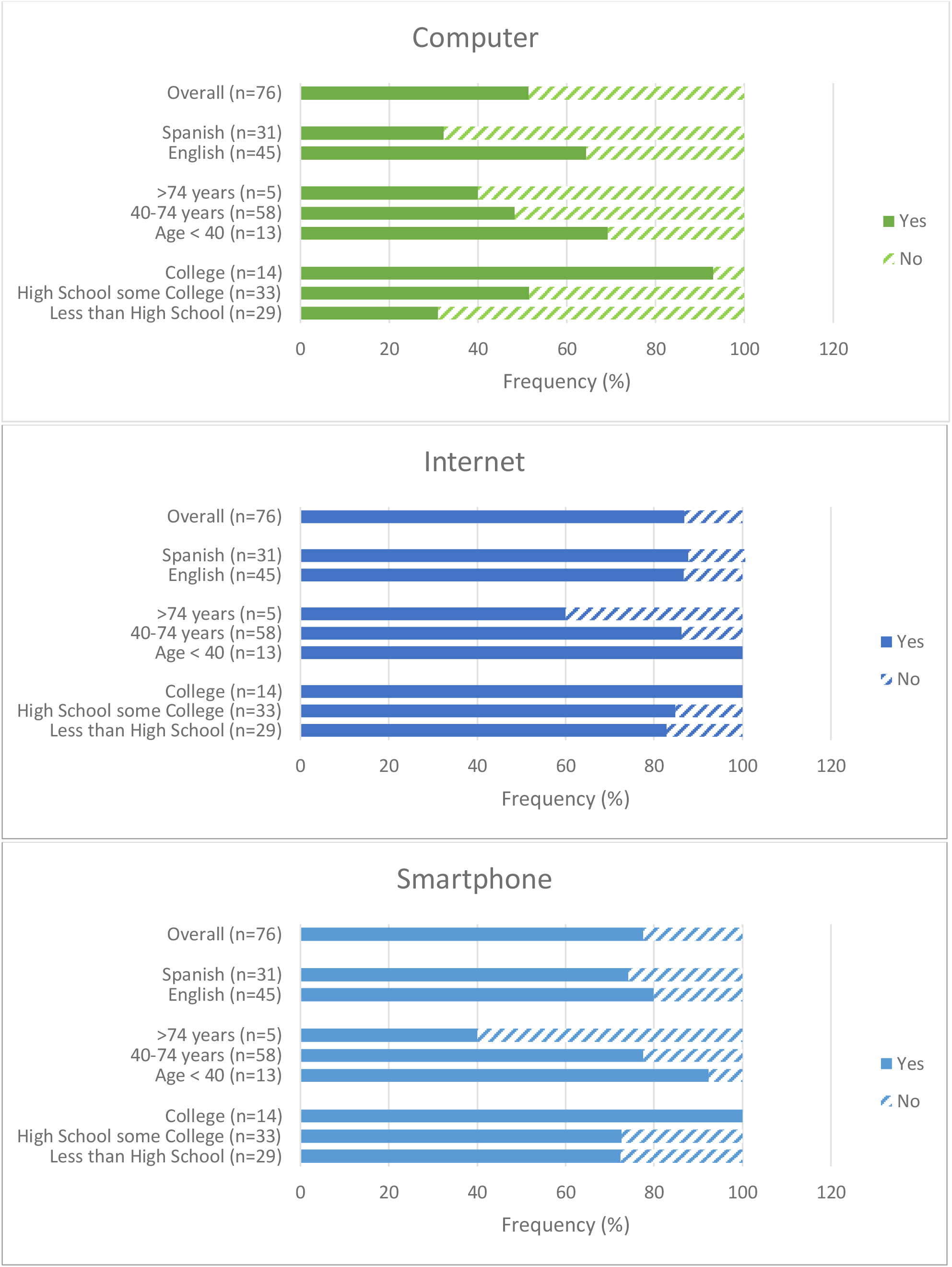
Access to digital tools overall and stratified by language, age, and educational level. Less than high school was defined as ‘No schooling, primary school, or some high school’. High school, Some college was defined as ‘High school graduate/diploma or the equivalent, some college/no degree, or trade/technical/vocational training’. College was defined as ‘associate, bachelors, masters or professional degree’.

Those with a high school degree and/or some college or trade school experience reported 51.4% access to computer, 83.8% access to internet, and 73% access to smartphones. Individuals with a college degree or more reported 93.8% access to computer, 93.8% access to internet, and 100% with access to a smartphone.

The digital literacy questionnaire was stratified and analyzed across age, education levels, and language (Figure 3, Table 2). Digital literacy was notably lower among Spanish speaking patients with ¾ of Spanish speakers reporting poor computer literacy and 60% reporting poor internet and smartphone literacy (p <0.001). Computer, internet, and smartphone literacy worsened with increased age. Less than 10% of patients 40 years of age or younger reported poor digital literacy for computer, smartphone, and internet use compared to 50%, 40% and 44% of patient 40-74 respectively, and 60% of patients over 74 years old (p= 0.002, 0.032, 0.023). A similar trend is seen among digital literacy and education level.

**Figure 3.**
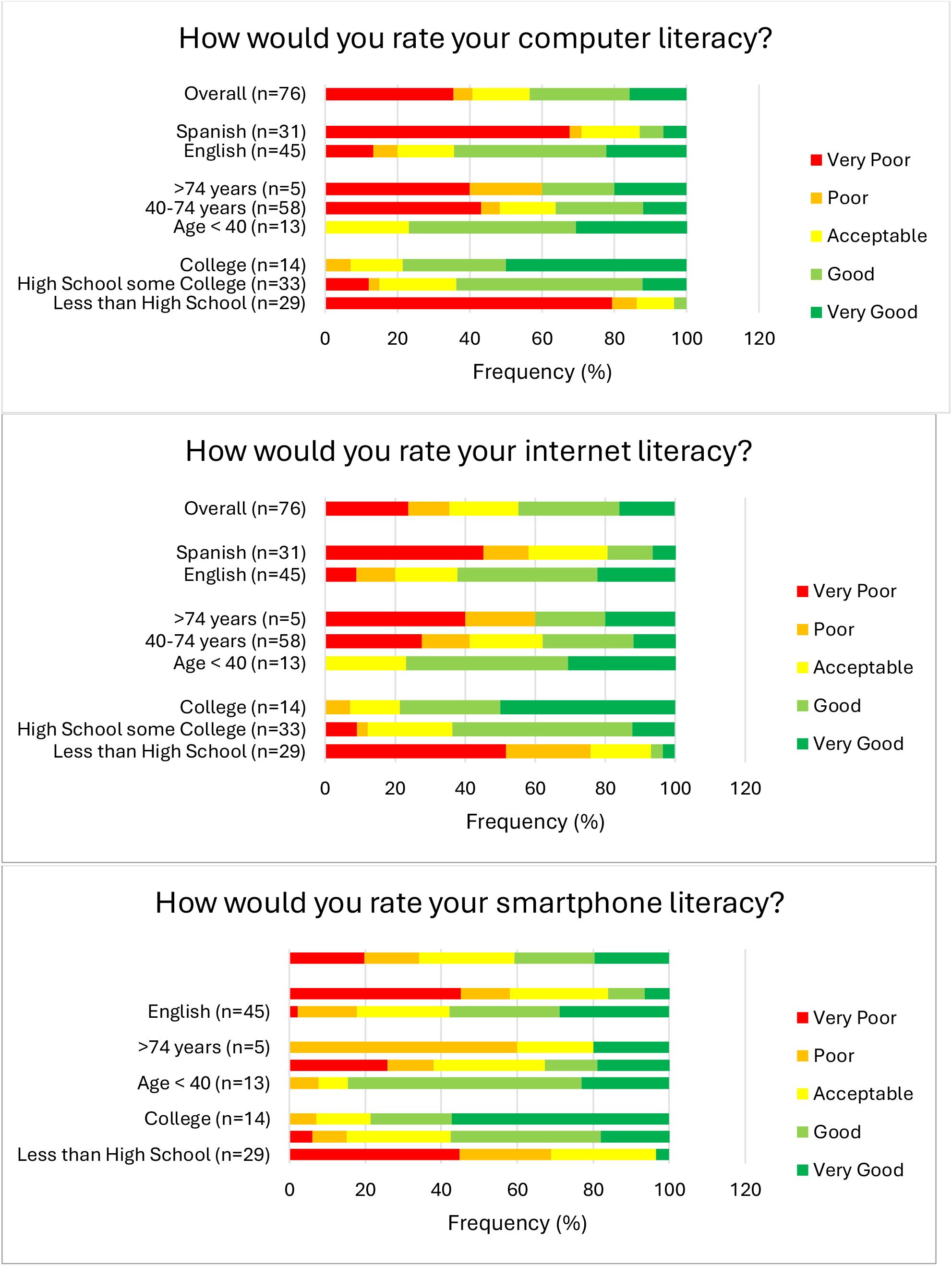
Digital literacy overall and stratified by educational level. Less than high school was defined as ‘No schooling, primary school, or some high school’. High school, Some college was defined as ‘High school graduate/diploma or the equivalent, some college/no degree, or trade/technical/vocational training’. College was defined as ‘associate, bachelors, masters or professional degree’.

Computer digital literacy is low among patients with less than high school education with over 85% of patients reporting poor or very poor computer literacy, 75% reporting poor or very poor internet literacy, and 70% reporting poor or very poor smartphone literacy (p<0.0001). This contrasted with high school educated individuals with about 15% reporting poor or very poor literacy for computer, internet, and smartphone, and college educated patients with 7% reporting poor literacy for computers and smartphones and 13% reporting poor internet literacy (p<0.001).

## Discussion

This study underscores the influence of sociodemographic factors such as language, age, and education level on patients’ health literacy, access to digital tools, and digital literacy in a breast surgical oncology clinic at an urban safety-net hospital (SNH). The findings align with and expand upon existing literature, emphasizing the ongoing disparities in healthcare access and outcomes among vulnerable populations.

### Health literacy

The validated health literacy questionnaire showed that patients who are Spanish speaking, older than 74 years of age, and/or with fewer years of education have poorer health literacy than their English speaking, younger, and more educated counterparts (Figure 1). These patients often require assistance with reading hospital materials, feel uncomfortable filling out medical forms independently, and struggle to understand written medical information. These findings align with previous studies indicating that lower health literacy is associated with poorer health outcomes, including increased emergency department visits, decreased medication adherence, and higher mortality rates among elderly patients^4^. The overestimation of patient literacy by healthcare providers and the reluctance of patients to disclose literacy challenges further complicate effective communication and care.

### Digital Literacy

Digital literacy disparities are evident across age, language, and education levels (Figure 3, Table 2). Older patients, particularly those over 75, show significantly reduced access to computers, the internet, and smartphones. Similarly, Spanish-speaking patients and those with less than a high school education report lower digital literacy and limited access to digital resources. This digital divide affects patients’ ability to engage with online health information and portals, which are increasingly essential for managing healthcare.

The lower digital literacy among non-English speakers, as well as those with limited education, exacerbates the challenges in accessing healthcare information and services. Previous studies have shown these populations are less likely to use digital tools^9^, which can lead to delays in care and poorer health outcomes. The correlation between higher income-to-needs ratio and increased internet usage^9^ suggests that economic factors play a crucial role in digital access and literacy as well.

### Implication for Clinical Practice

The disparities identified in this study necessitate targeted interventions to improve health and digital literacy among vulnerable populations.

1. Enhanced Patient Education:
  - Implementing tailored educational programs that address the specific needs of low-literacy and non-English-speaking patients. These programs should use clear, simple language and culturally appropriate materials.
  - Training healthcare providers to recognize and address literacy challenges, ensuring that communication is effective and comprehensible.
2. Improved Access to Digital Resources:
  - Expanding access to digital tools and the internet for underserved populations. This can include providing computers and internet access in community centers and healthcare facilities.
  - Offering digital literacy training programs to help patients navigate online health information and portals.
3. Supportive Services:
  - Increasing the availability of medical interpreters and culturally competent care coordinators to assist non-English-speaking patients.
  - Developing partnerships with community organizations to provide support and resources for patients facing multiple barriers to care.

### Limitations

One limitation of this study is the surveyed population size. Response rates among those greater than 74 years old and those less than 40 years old were less than 5 in some categories. This can be partially attributed to the specific population sampled, given the low numbers of patients diagnosed with breast cancer within either of those ranges. Additionally safety net hospitals, like the one in this study, often serve a smaller proportion of older adults than non-safety net hospitals ^16^.

### Future Research

Further research is needed to explore the long-term impact of improved health and digital literacy on patient outcomes. Longitudinal studies could assess how targeted interventions influence healthcare utilization, adherence to treatment, and overall health status. Additionally, investigating the specific barriers faced by different demographic groups can inform the development of more precise and effective strategies to bridge the literacy gaps.

## Conclusion

The study demonstrates the critical need to address health and digital literacy disparities to ensure equitable care for breast cancer patients at SNHs. By recognizing and addressing the sociodemographic factors that influence patient outcomes, healthcare providers can implement targeted interventions to improve communication, access to care, and overall health outcomes for vulnerable populations.

## Data Availability

All data produced in the present study are available upon reasonable request to the authors

## Appendix A Breast Clinic Survey

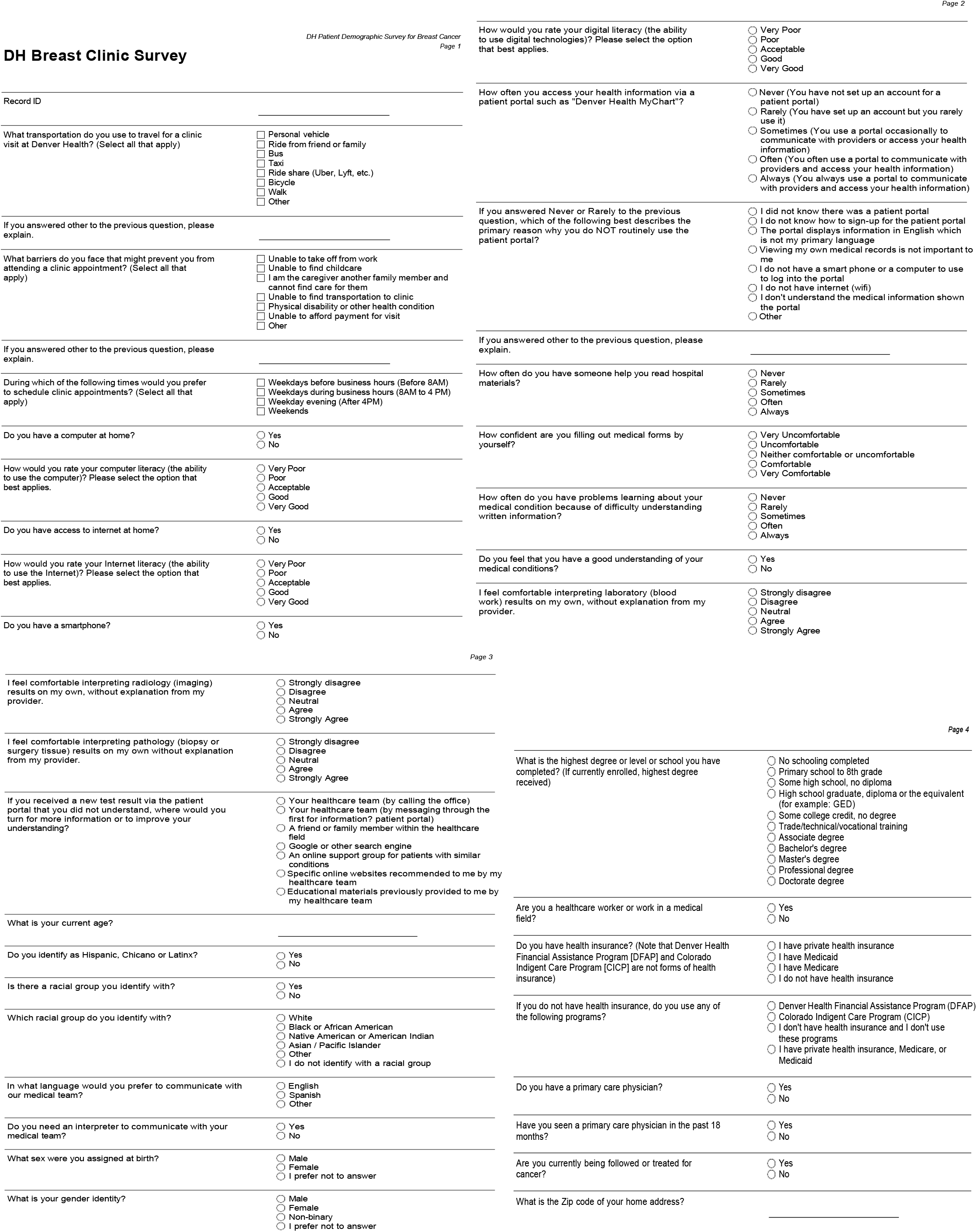

|  |  |
| --- | --- |
| Record ID |  |
| What transportation do you use to travel for a clinic visit at Denver Health? (Select all that apply) | ☐ Personal vehicle<br>☐ Ride from friend or family<br>☐ Bus<br>☐ Taxi<br>☐ Ride share (Uber, Lyft, etc.)<br>☐ Bicycle<br>☐ Walk<br>☐ Other |
| If you answered other to the previous question, please explain. |  |
| What barriers do you face that might prevent you from attending a clinic appointment? (Select all that apply) | ☐ Unable to take off from work<br>☐ Unable to find childcare<br>☐ I am the caregiver another family member and cannot find care for them<br>☐ Unable to find transportation to clinic<br>☐ Physical disability or other health condition<br>☐ Unable to afford payment for visit<br>☐ Other |
| If you answered other to the previous question, please explain. |  |
| During which of the following times would you prefer to schedule clinic appointments? (Select all that apply) | ☐ Weekdays before business hours (Before 8AM)<br>☐ Weekdays during business hours (8AM to 4 PM)<br>☐ Weekday evening (After 4PM)<br>☐ Weekends |
| Do you have a computer at home? | ☐ Yes<br>☐ No |
| How would you rate your computer literacy (the ability to use the computer)? Please select the option that best applies. | ☐ Very Poor<br>☐ Poor<br>☐ Acceptable<br>☐ Good<br>☐ Very Good |
| Do you have access to internet at home? | ☐ Yes<br>☐ No |
| How would you rate your Internet literacy (the ability to use the Internet)? Please select the option that best applies. | ☐ Very Poor<br>☐ Poor<br>☐ Acceptable<br>☐ Good<br>☐ Very Good |
| Do you have a smartphone? | ☐ Yes<br>☐ No |
| How would you rate your digital literacy (the ability to use digital technologies)? Please select the option that best applies. | ☐ Very Poor<br>☐ Poor<br>☐ Acceptable<br>☐ Good<br>☐ Very Good |
| How often you access your health information via a patient portal such as "Denver Health MyChart"? | ☐ Never (You have not set up an account for a patient portal)<br>☐ Rarely (You have set up an account but you rarely use it)<br>☐ Sometimes (You use a portal occasionally to communicate with providers or access your health information)<br>☐ Often (You often use a portal to communicate with providers and access your health information)<br>☐ Always (You always use a portal to communicate with providers and access your health information) |
| If you answered Never or Rarely to the previous question, which of the following best describes the primary reason why you do NOT routinely use the patient portal? | ☐ I did not know there was a patient portal<br>☐ I do not know how to sign-up for the patient portal<br>☐ The portal displays information in English which is not my primary language<br>☐ Viewing my own medical records is not important to me<br>☐ I do not have a smart phone or a computer to use to log into the portal<br>☐ I do not have internet (wifi)<br>☐ I don't understand the medical information shown the portal<br>☐ Other |
| If you answered other to the previous question, please explain. |  |
| How often do you have someone help you read hospital materials? | ☐ Never<br>☐ Rarely<br>☐ Sometimes<br>☐ Often<br>☐ Always |
| How confident are you filling out medical forms by yourself? | ☐ Very Uncomfortable<br>☐ Uncomfortable<br>☐ Neither comfortable or uncomfortable<br>☐ Comfortable<br>☐ Very Comfortable |
| How often do you have problems learning about your medical condition because of difficulty understanding written information? | ☐ Never<br>☐ Rarely<br>☐ Sometimes<br>☐ Often<br>☐ Always |
| Do you feel that you have a good understanding of your medical conditions? | ☐ Yes<br>☐ No |
| I feel comfortable interpreting laboratory (blood work) results on my own, without explanation from my provider. | ☐ Strongly disagree<br>☐ Disagree<br>☐ Neutral<br>☐ Agree<br>☐ Strongly Agree |
| I feel comfortable interpreting radiology (imaging) results on my own, without explanation from my provider. | ☐ Strongly disagree<br>☐ Disagree<br>☐ Neutral<br>☐ Agree<br>☐ Strongly Agree |
| I feel comfortable interpreting pathology (biopsy or surgery tissue) results on my own without explanation from my provider. | ☐ Strongly disagree<br>☐ Disagree<br>☐ Neutral<br>☐ Agree<br>☐ Strongly Agree |
| If you received a new test result via the patient portal that you did not understand, where would you turn for more information or to improve your understanding? | ☐ Your healthcare team (by calling the office)<br>☐ Your healthcare team (by messaging through the first for information? patient portal)<br>☐ A friend or family member within the healthcare field<br>☐ Google or other search engine<br>☐ An online support group for patients with similar conditions<br>☐ Specific online websites recommended to me by my healthcare team<br>☐ Educational materials previously provided to me by my healthcare team |
| What is your current age? |  |
| Do you identify as Hispanic, Chicano or Latinx? | ☐ Yes<br>☐ No |
| Is there a racial group you identify with? | ☐ Yes<br>☐ No |
| Which racial group do you identify with? | ☐ White<br>☐ Black or African American<br>☐ Native American or American Indian<br>☐ Asian / Pacific Islander<br>☐ Other<br>☐ I do not identify with a racial group |
| In what language would you prefer to communicate with our medical team? | ☐ English<br>☐ Spanish<br>☐ Other |
| Do you need an interpreter to communicate with your medical team? | ☐ Yes<br>☐ No |
| What sex were you assigned at birth? | ☐ Male<br>☐ Female<br>☐ I prefer not to answer |
| What is your gender identity? | ☐ Male<br>☐ Female<br>☐ Non-binary<br>☐ I prefer not to answer |
| What is the highest degree or level or school you have completed? (If currently enrolled, highest degree received) | ☐ No schooling completed<br>☐ Primary school to 8th grade<br>☐ Some high school, no diploma<br>☐ High school graduate, diploma or the equivalent (for example: GED)<br>☐ Some college credit, no degree<br>☐ Trade/technical/vocational training<br>☐ Associate degree<br>☐ Bachelor's degree<br>☐ Master's degree<br>☐ Professional degree<br>☐ Doctorate degree |
| Are you a healthcare worker or work in a medical field? | ☐ Yes<br>☐ No |
| Do you have health insurance? (Note that Denver Health Financial Assistance Program [DFAP] and Colorado Indigent Care Program [CICP] are not forms of health insurance) | ☐ I have private health insurance<br>☐ I have Medicaid<br>☐ I have Medicare<br>☐ I do not have health insurance |
| If you do not have health insurance, do you use any of the following programs? | ☐ Denver Health Financial Assistance Program (DFAP)<br>☐ Colorado Indigent Care Program (CICP)<br>☐ I don't have health insurance and I don't use these programs<br>☐ I have private health insurance, Medicare, or Medicaid |
| Do you have a primary care physician? | ☐ Yes<br>☐ No |
| Have you seen a primary care physician in the past 18 months? | ☐ Yes<br>☐ No |
| Are you currently being followed or treated for cancer? | ☐ Yes<br>☐ No |
| What is the Zip code of your home address? |  |

